# Uncovering the Dual Role of Mitochondrial and Nuclear DNA Variants in Pediatric Cardiomyopathies

**DOI:** 10.1101/2023.12.20.23300082

**Authors:** M. Arda Temena, Ebru Erzurumluoglu Gokalp, Ezgi Susam, Duygu Cinar, Hikmet Kiztanir, Pelin Kosger, Beyhan Durak Aras, Sevilhan Artan, Oguz Cilingir

**Affiliations:** Science & Technology Policy Studies, METU, Turkey; Medical Genetics Department, ESOGU Faculty of Medicine, Turkey; Child Health and Diseases Department, Faculty of Medicine, ESOGU Faculty of Medicine, Turkey

**Author notes:** **Correspondence:** Mehmet Arda Temena METU, Science and Technology Policy Studies, Ankara, Turkey, Mail.

## Abstract

Myocardial defects, originating from disruptions in genes affecting mitochondrial proteins interacting with others, have received limited research attention. In our study involving 27 pediatric cardiomyopathy patients, we explored mitochondrial and nuclear genes for four main cardiomyopathy subtypes. Sequencing cardiomyopathy-associated genes in patients was followed by whole mtDNA sequencing in these individuals, with 31 healthy pediatric controls for the latter part. Our findings uncovered significant pathogenic variants: MYH7 variants in three hypertrophic cardiomyopathy cases, a notable TNNC1 variant inherited interestingly in an autosomal recessive manner in two related restrictive cardiomyopathy patients, and a pathogenic TRDN variant in a left-ventricular non-compaction patient. Both a variant in FOXRED1, functioning in mitochondrial complex I stability, and a MT-CO1 variant were detected in two siblings, influencing early onset together. Additionally, a novel MT-RNR1 variant (m.684T>C) in one case might explain the phenotype. Our study highlights how both mtDNA and nDNA variants could have interconnected roles in understanding cardiomyopathy genetics. This study emphasizes that the functional studies are needed to recognize this dual relationship within bioenergetic pathways in cardiac muscle.

## INTRODUCTION

In pediatric population, heart failure can occur when normal myocardium is mostly damaged by an inflammatory or infectious process like myocarditis, metabolic disorders that cause cardiomyopathy, and solely due to hemodynamic imbalances because of syndromic or non-syndromic congenital structural defects(Iaizzo, 2015). Although cardiomyopathies one of the leading causes of heart failure and sudden cardiac death among the young all over the world, present molecular studies are not yet to be concluded and therapeutic approaches are used mostly to relieve the symptoms of the disease except heart transplantation at the end stage of the disease(Lipshultz et al., 2019; Sanganalmath & Bolli, 2013). Therefore, pediatric cardiomyopathies require an elaborative and multidisciplinary evaluation approach starting from diagnosis to prevent rapid progressive case of the disease(Lee et al., 2017).

The prevalence of pediatric cardiomyopathy subtypes (classification made by American Heart Association, European Society of Cardiology and World Heart Federation(Elliott et al., 2007; B. Maron et al., 2006),(Arbustini et al., 2014)) differs based on the accuracy of diagnostic evaluations and classification criteria, and the rarity of the disease complicates data collection. Indeed, it has been reported that certain genotypes may correspond to specific types of cardiomyopathies(J. W. McKenna et al., 2017),(Tobita et al., 2018).

It is known that some variants in some certain genes known to be directly associated with cardiomyopathies may have non-cardiac systemic or metabolic manifestations(Fiorillo et al., 2016; Jhang et al., 2016). Given the complexity of genetic factors involved, there is limited but growing understanding of the portion of heritability that remains unexplained interactions between genes, and factors that modify the progression of each type of cardiomyopathy(Lee et al., 2017). Pediatric cardiomyopathies can result from various genetic alterations, including metabolic dysfunctions like a diabetic mother and the occurrence of hypertrophic cardiomyopathy-HCM in some storage disorders. The genetic basis for isolated pediatric cardiomyopathies may overlap with those found in adult cases, but the mode of inheritance can be variable and may also occur due to solely *de novo* mutations(Jhang et al., 2016; Lee et al., 2017). Recent research has shown that genetic factors adapting to various inheritance patterns have a significant role in cardiomyopathy which helps to determine a correct classification(Hershberger et al., 2013; Jacoby & McKenna, 2011). Among those, although much smaller than the nuclear genome, studies on the mitochondrial genome also have become the most popular subject not only for cardiomyopathies and particularly for cardiomyocytes(Dorn, 2015) but also for investigating other complex diseases(Tashiro, 2018).

In this study, we sought to investigate the genetic profile of pediatric patients with primary cardiomyopathy from Turkey’s Central Anatolia Region by sequencing cardiomyopathy-related genes and whole mtDNA sequencing. **Figure 1** provides a graphical abstract illustrating the design of the study and significant results. There is a graphical abstract for this study. Additionally, the aim was to assess patients in the primary cardiomyopathy group given the crucial role that mitochondrial proteins play in energy metabolism and bioenergetics in relation to the heart along with nuclear DNA (nDNA) variants.

**Figure 1.**
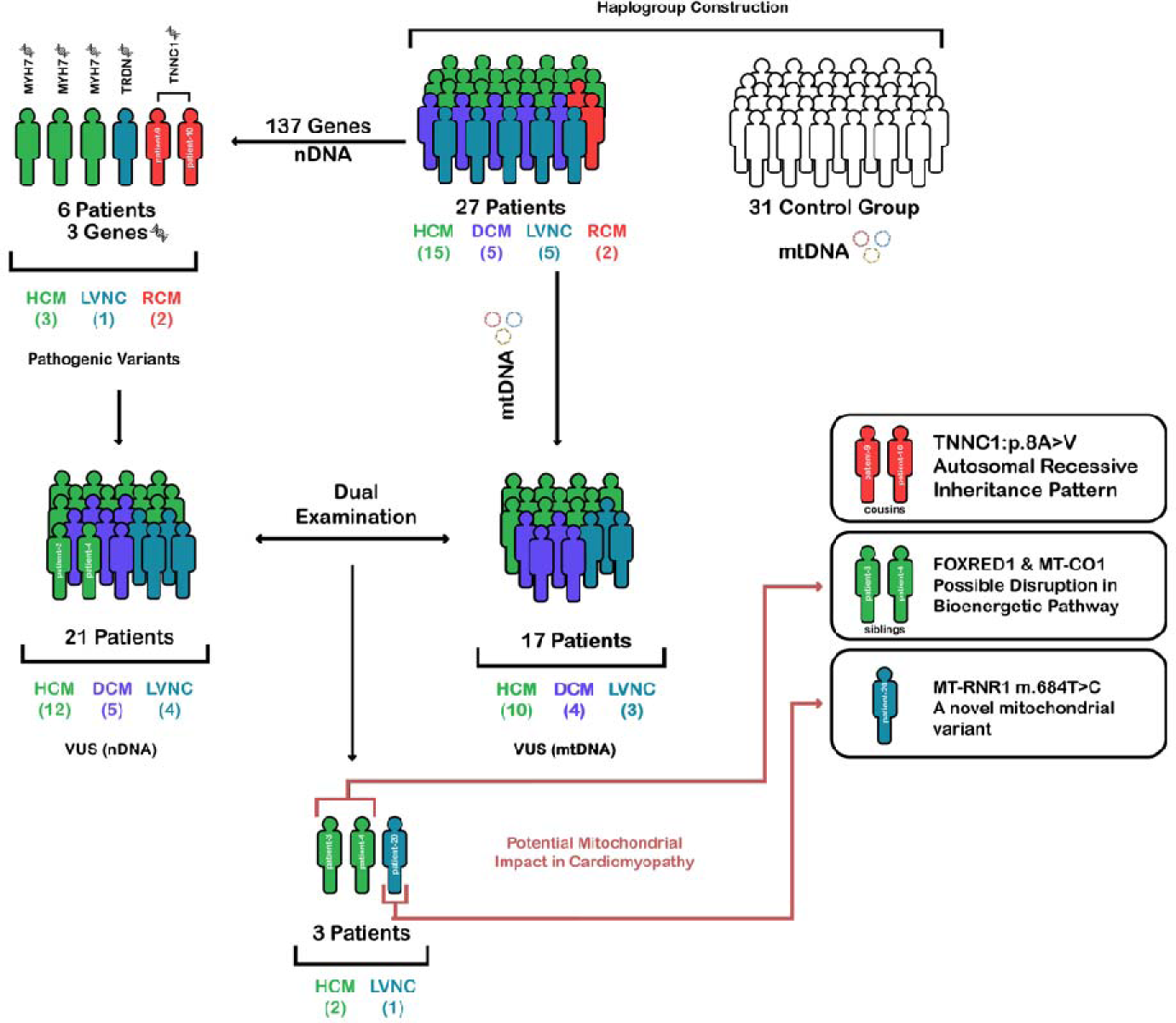
Genetic profiling in pediatric cardiomyopathies. This graphical abstract illustrating the comprehensive genetic profiling approach used in this study to sequence cardiomyopathy-related genes and whole mitochondrial DNA in pediatric patients with cardiomyopathy.

## RESULTS

### Clinical evaluation

All detailed clinical manifestations and other related information are analyzed and will be available with **Supplementary Table S1** upon reasonable request to corresponding author.

### Cardiomyopathy-associated variants

A total of 269 exonic variants were identified after filtering (**Figure 2B**). Out of these variants, 164 (60.97%) were nonsynonymous SNVs whereas 98 of them (36.43%) were synonymous. There were also three frameshift deletions (1.12%) and three non-frameshift deletions (1.12%), and one stop-gain variant (0.37%). Following the variants are classified by gene, and the count and percentage of variants found in each gene are given, the total number of variants is 269, of which 12 (4.5%) are homozygous and 257 (95.5%) are heterozygous. The most frequent variants are found in the gene TTN with 91 (33.8%) variants and the second one is in PLEC gene with 24 (8.9%). A scatterplot that summarizes the variants grouped by genes and corresponding genotypes was given in **Supplementary Figure S1**. The expression levels of these genes in the heart were examined by the GTEx database, and the resulting plot is presented in **Figure 2C**. This analysis revealed four clusters for the genes in which some variants detected and among them, MYH7, MYBP3, TNNT2 and TNNC1 is highly expressed.

**Figure 2.**
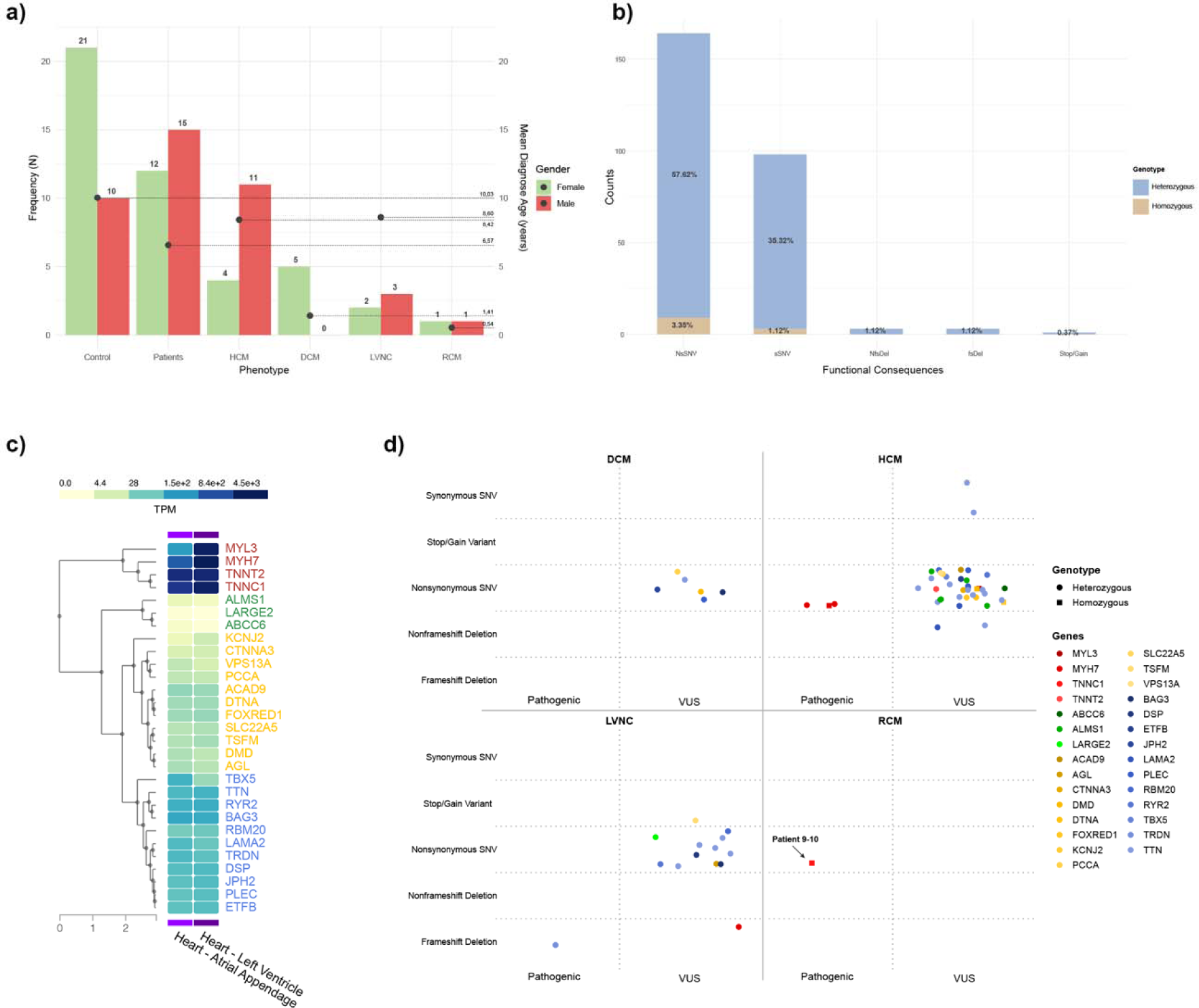
General demographic features of the patients and the study highlights. **A.** Distribution of number of patients and control group, and mean age (for mtDNA group) over cardiomyopathy subtypes There are four distinct primary cardiomyopathy group (HCM; hypertrophic cardiomyopathy, DCM; dilated cardiomtyopathy, RCM; restrictive cardiomyopathy, LVNC; left ventricular non compactiomn). **B.** Functional consequences of nDNA variants from patients’ group is given accros their genotype of heterozygosity or homozygosity. **C.** Expression level of VUS and pathogenic variant-detected genes in heart tissue. The graph was generated using Genotype-Tissue Expression (GTEx) website. The genes were colored according to their expression level clusters (Very high expression; red, high expression; blues, moderate expression; yellow, low expression; green). **D.** Representation of VUS and pathogenic variants over disease subtypes and genotypes. Each gene was colored according to their corresponding expression clusters.

In the cohort of 27 pediatric patients with cardiomyopathy, 62 variants were identified after evaluation and filtering as VUS or Pathogenic. All these variants grouped by disease subtype can be found in **Figure 2D**. Out of these variants, 57 (91.9%) were heterozygous and five of them (8.1%) were homozygous. Most variants were nonsynonymous SNVs, 55 (88.7%) of which 5 were homozygous. There were also 2 variants each of frameshift deletions (3.2%) and non-frameshift deletions (3.2%), and one stop-gain variant (1.6%). Two variants were Synonymous SNV (3.2%). Out of 62 variants, 5 (8.1%) were pathogenic, of which 3 were nonsynonymous SNV in MYH7 gene leading to HCM, 1 was nonsynonymous SNV in TNNC1 leading to RCM, 1 was frame shift deletion in TRDN leading to LVNC. The remaining 57 variants (91.9%) were concluded as VUS.

Specifically, patient-8, patient-13, and patient-14 have a pathogenic variant in the MYH7 gene, and they are diagnosed with HCM (**Supplementary Figure S2**). Patient-21 has a pathogenic variant in the TRDN gene and patient 9-10, who were mother side cousins and diagnosed with RCM have a pathogenic variant in the TNNC1 (**Supplementary Figure S2**).

### Mitochondrial variants

Results show that in a cohort of 27 pediatric patients with cardiomyopathy, the analysis of mitochondrial variants revealed that 695 alterations were found in the cardiomyopathy (CM) group, while 315 unique alterations were found in the control group. Examination of different disease groups showed that the highest frequency was found in HCM, with 337 variants, followed by LVNC with 164 variants, DCM with 133 variants, and RCM with 61 variants. On the other hand, ratio of variants over patients per disease subgroup are DCM at 26.6, HCM at 22.0, LVNC at 31.8 and RCM at 30.5. Analysis of the genetic alterations revealed that the most common type of alteration was SNPs, with 677, followed by insertions (INS) with 10, multi-nucleotide polymorphism (MNP) with 3, and Deletion (DEL) with 5 occurrences. In **Figure 3**, the distribution of all variants among the different disease subtypes is shown in the patient group. Additionally, all variants found in patients with control group and only control group were also plotted (**Supplementary Figure S3 and S4**, respectively). However, we evaluated variants that are found only in patient group after comparison with control group.

**Figure 3.**
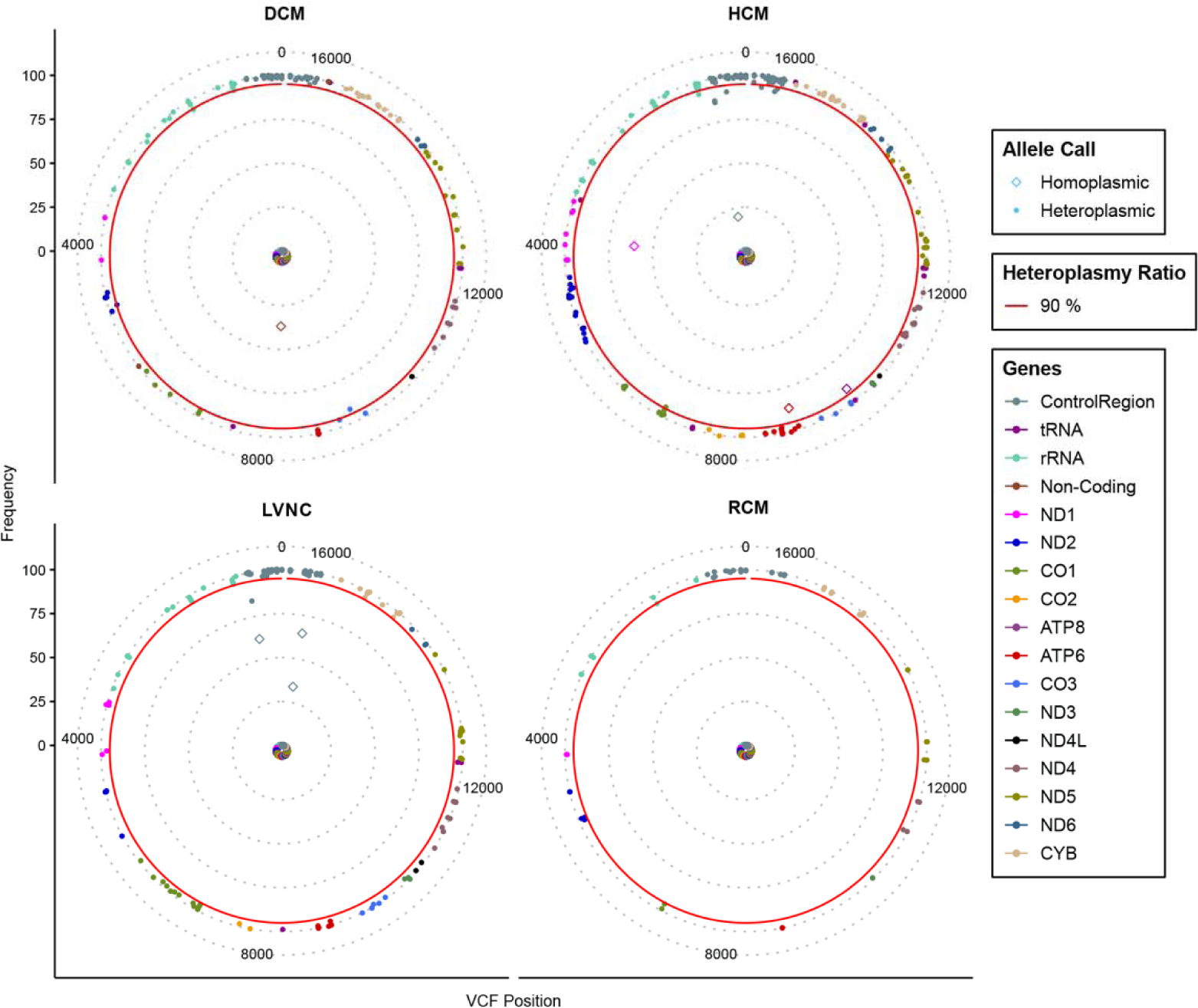
Solar Plot of Mitochondrial Variants in Cardiomyopathy Subtypes. Mitochondrial variants across cardiomyopathy subtypes from mtDNA sequencing are displayed on a solar plot. An identified variant is represented by each dot. According to the plot legend, the inner dashed rings of the plot represent the level of heteroplasmy, and dots are colored according to genes at that region. The position of the mitochondrial base pair is shown on the X-axis. Red line represents a line for specified 90% heteroplasmy level.

Results showed that homoplasmic variants were the most common, with 688, while heteroplasmic variants were found in 7 occurrences. Seven variants detected in only five patients showed heteroplasmic features. These are m.16184C>T detected in patient-16 (34%), m.8286T>C detected in patient-18 (37.6%), m.3918G>A detected in patient-22 (30%), m. 9053G>A and m.10045T>C (87.5% and 92.3%, respectively) and m.16189Tdel (66.1%) detected in patient-26. All other variants detected in mtDNA show homoplasmic (≥95%) feature. After filtering of all variants, mtDNA sequencing results revealed that 34 transition were found, one insertion was detected. In **Figure 4A**, functional consequences and disease subtypes of filtered mtDNA variants were given. The mitochondrial loci were examined, revealing a total of 33 unique variants (35 in total) across two rRNA and ten protein coding genes. The 16S locus had the highest number of alterations with six variants, followed by COI with five variants, 12S and ND5 with four variants each. Three genes had three variants each: ATPase6, COIII, and Cytb. Two genes had two variants each: ND4, and ND6. Finally, three genes had only one variant each: ND2, ND3, and ND4L. Also, the mitochondrial haplogroups of patients and controls in the study were analyzed (**Figure 4B**). The most common major haplogroups among patients were H, J, and U while the most common haplogroups among controls were H and U. There was no statistically significant consequence between control and patient group with respect to haplogroups. Other major haplogroups are Y, W, K, M, V, T, X, R, C and N. Additionally, another plot (**Supplementary Figure S5**) was provided where mitochondrial variants were classified according to the patients.

**Figure 4.**
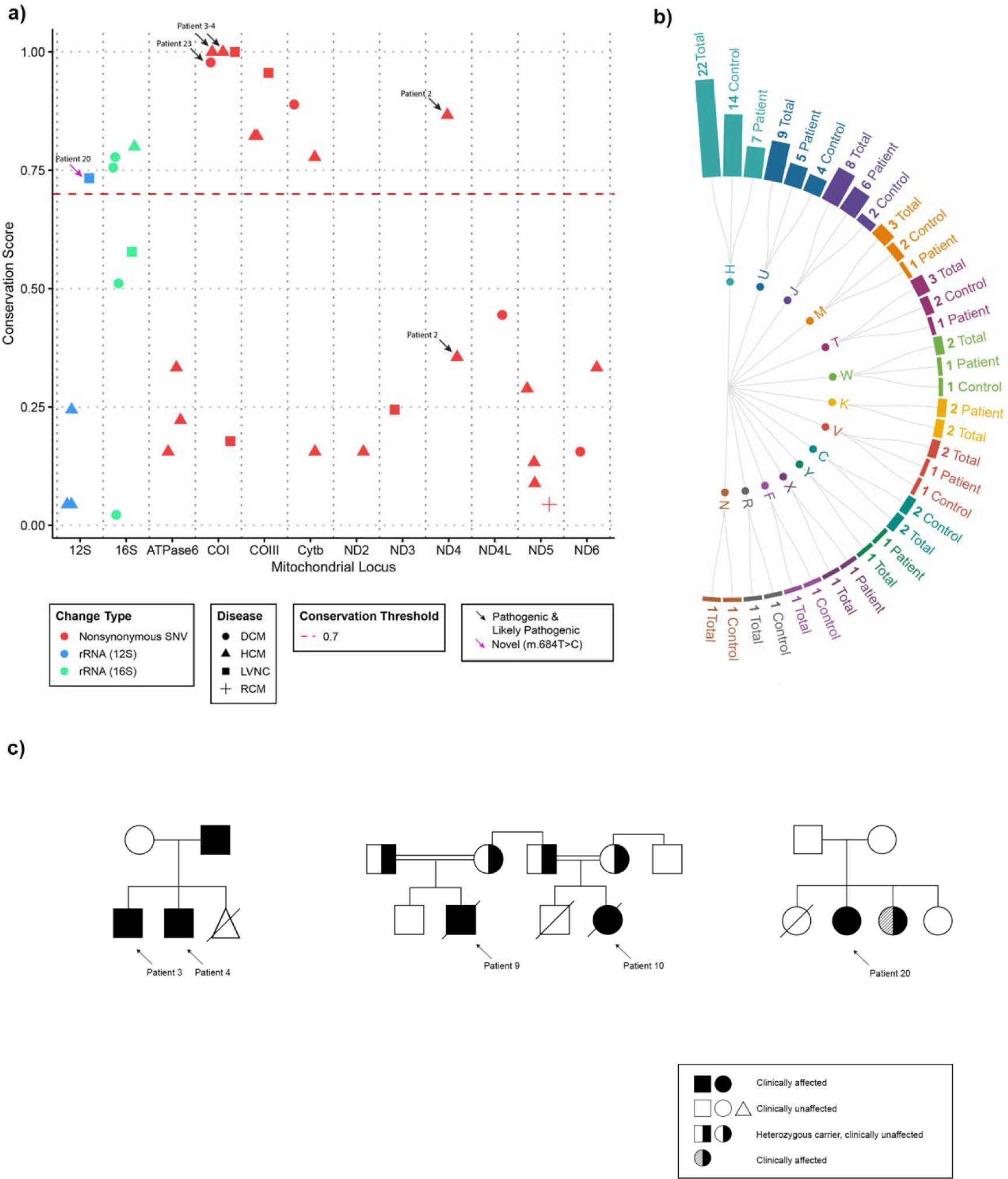
Overall identified and filtered mtDNA Variants and Pedigree analysis of the patients. **A.** Filtered variants across mtDNA genes and functional consequences of these variants. Alteration type and the subtype of the patient determines the color and the shape, respectively. Conservation score line was given as red dashed line. Variants that are indicated by arrows are either pathogenic or likely pathogenic which are depicted as black or pink color, respectively. **B.** mtDNA haplogroups distribution for the study cohort. **C.** Three pedigrees are given for patients, but detailed pedigrees are available on request to corresponding authors.

Haplogroup subtypes as well as a comprehensive list of all filtered mtDNA and nDNA variants with their pathogenicity predictions were provided in **Supplementary Table S2A, Table S2B,** and **Table S2C**, respectively, but patient information is removed and available on request.

## DISCUSSION

Looking at the distribution of clinical and demographic characteristics among the patients, we see parallel results with a comprehensive study conducted among pediatric cardiomyopathy patients in 2022(Ware et al., 2022). Although the 27% positive family history rate (**Supplementary Table S1A**) among our patients may not directly correspond to the 36% rate in that study, this could be due to the filters used in our study and the low number of patients in our study. In addition, the age of diagnosis was found to be high on average in our patient group. While this could be due to asymptomatic patients receiving a diagnosis only during routine check-ups or other illnesses, socioeconomic factors that affect the access of young children/families to healthcare services in the country should not be ignored. Moreover, one another comprehensive study revealed positive association between cardiomyopathy subtype and age at diagnosis(Bagnall et al., 2022). Similarly, the diagnosis of HCM was made at later ages compared to other subtypes, while in cases of DCM, it was made before the age of 3 in our study. However, the low number of patients included in the study and the rarity of the disease may be a reason for the discrepancy in the mean age of diagnosis for LVNC and RCM subtypes.

### Hypertrophic cardiomyopathy

Out of 15 HCM patients, either a VUS or pathogenic variant was detected in all of them. In three HCM patients (20% of HCM patients), a pathogenic variant was detected in the MYH7 gene, which is consistent with the information that the variants found in HCM children are most commonly in the MYH7 gene(Ware et al., 2022). Among them, patient-13 and patient-14 was heterozygous for a nonsynonymous single nucleotide variant (SNV) in the MYH7 gene as p.Arg403Gln and p.Arg719Trp, respectively. Additionally, patient-8 was found to be homozygous for a nonsynonymous SNV in the MYH7 gene (p.Asp928Val). In fact, MYH7 mutations can lead to a direct effect of delayed relaxation due to abnormal myosin function at the molecular level, which in turn causes inefficient energy use at the myocardial tissue level(Alamo et al., 2017; Nguyen et al., 2022; Toepfer et al., 2020). In patients with MYH7, the most common symptom reported was palpitations in one study(Velicki et al., 2020) but other symptoms of HCM are fatigue, chest pain, and syncope(B. J. Maron, 2018). These patients were assigned to different mitochondrial haplogroups, H, U and W respectively.

In addition to the MYH7 variants mentioned, all variants identified in patients with HCM have been classified as variants of unknown significance (VUS). A previous functional study of a missense mutation in the MYL3 gene, previously classified as VUS, suggested that the mutation had benign effects(H. Guo et al., 2021). However, a homozygous alteration (MYL3(NM_000258.3):c.170C>A/p.Ala57Asp) with damaging prediction results and the high expression of the gene in left ventricular (**Figure 2C**), and the presence of highly conserved a mitochondrial variant in MT-RNR2 (m.2060A>G) in the patient-11 may possibly contribute to the phenotype altogether. Moreover, a missense variant in TNNT2 gene is identified in patient-17 with asymmetric septal hypertrophy (TNNT2(NM_001276345.2):c.691A>G/p.Ile231Val). It affects the TNNT2 gene, which codes for cardiac troponin T (cTnT), a protein that is part of the troponin complex found in heart muscle and plays an important role in regulating calcium as well as thin actin filament contractility and expressed highly in left ventricle (**Figure 2C**). Apart from this variation, this patient has other missense variants in TTN, PLEC and LAMA2 gene as well as in MT-ND6 and MT-CYB gene latter of which has high conservation rate (78%). One study by Rippol-Vera et al. suggests that some TNNT2 variants may interact with other variants associated with other cardiac diseases or pathways, influencing the way the disease develops and the prognosis(Ripoll-Vera et al., 2016). This suggests that further studies are needed to confirm this, and this patient’s variants could be a fine model for a functional study.

Furthermore, there are some publications linking RBM20 to cardiomyopathy(W. Guo et al., 2012; Lennermann et al., 2020). Additionally, a study by Zaragoza et al. in 2011, which examined the relationship between mtDNA and cardiomyopathy, classified a variant on MT-ND4 as highly likely to be pathogenic(Zaragoza et al., 2011). In our study, when *in silico* analysis scores for pathogenicity were considered for the variant with its high conservation score on MT-ND4 (m.11084A>G), it is possible that this variant may have created a pathogenic situation in patient-2, in combination with RBM20 variant (RBM20(NM_001134363.3):c.1745A>T / p.Asn582Ile). Next, patient-3 and patient-4 are siblings who have been diagnosed with HCM. Both patients were found to have heterozygous variants in the TTN gene, which is known to be associated with HCM. Additionally, patient-3 and-4 were also found to have heterozygous missense variant in the FOXRED1 (FOXRED1(NM_017547.4:c.658C>T)/p.Pro220Ser). Some other variants in ALMS1 and ABCC6 genes, which were not directly related with primary cardiomyopathy though, were found in patient-4 whereas patient-3 only has additional TTN variants. Therefore, it is not clear how these variants are contributing to the development of HCM in these patients and further research is needed to understand the relationship between these genes and HCM. On the other hand, these siblings also have highly conserved mitochondrial variants in MT-CO1, concluded as pathogenic *in silico* analysis (m.6345T>C) and MT-CO3 (m.9966G>A) gene, with the conservation score of 100% and 82%, respectively. The cytochrome c oxidase, also known as Complex-IV, is the enzyme in the electron transport chain that transfers electrons from cytochrome c to molecular oxygen. It is made up of 14 subunits in humans, but only two of them, MT-CO1 and MT-CO2, have catalytic activity but MT-CO3 do not have this function. Also, one another mitochondrial gene FOXRED1, found in the list of mitochondrial proteins in Mitocarta 3.0(Rath et al., 2021), some variants of which is associated with mitochondrial Complex-I deficiency and related to cardiomyopathy(Mukherjee & Ghosh, 2020), encodes structural subunits and assembly factors. Co-existence of these two variants in FOXRED1 and MT-CO1 could explain the early-onset etiology of HCM in these siblings whose father has also has late-onset HCM and pedigree is given in **Figure 4C** as some variants inherited together can cause diverse group of mitochondrial diseases(Fernandez-Vizarra & Zeviani, 2021; Vercellino & Sazanov, 2022). Identified variants in MT-CO1 and FOXRED1 genes can be associated with early-onset cardiomyopathy, emphasizing their relevance within mitochondrial diseases in children, especially in the context of complex I disorders (Koopman et al., 2016).

Although recent studies revealed that DTNA variants were commonly found in patients with the left ventricular trabeculation and non-compaction phenotype which are also some manifestations of Barth syndrome(Bottillo et al., 2016; Geske et al., 2018; Moric-Janiszewska & Markiewicz-Łoskot, 2008), they were not sufficient to fully explain the clinical presentation of patient-1 (DTNA(NM_001390.4):c.165A>G/p.Ile55Met) with HCM in addition to presence of a TTN variant. In patient-12, variants in the CTTNA3, TBX5 and PLEC genes were classified as VUS. Also, a missense variant in the alpha-T-catenin gene (CTNNA3) is also identified, which plays a role in how cells stick together and is present in both the heart and brain. This variation may be linked to a type of heart disease called arrhythmogenic cardiomyopathy, but there is no proof that it is directly associated with HCM (B. Li et al., 2020). However, TBX which specifically plays an important role in cardiac contraction (Reactome ID: R-HSA-5576891.1) and the patient’s other variant on PLEC, there may be an explanation for the patient’s phenotype if a functional study is conducted harboring these mutations even though TBX5 and PLEC are not directly involved in primary cardiomyopathies (OMIM ID: 601282 and 601620). In addition, patient-22, has five variants, three of which are found in the TTN gene. The remaining two variants are in the DMD and RYR2 genes, and it is not clear how they contribute to the patient’s condition without further study of the patient’s family. Similarly, patient-25 also has three VUS located in the TTN, ALMS1, which is not directly involved in primary isolated cardiomyopathies, and SLC22A5 genes, all of which are missense mutations. Lastly, patient-6 has a homozygous nonsynonymous SNV in the KCNJ2 gene. Screening the KCNJ2 gene in individuals is clinically significant in syndromic cases because a significant proportion of individuals with pathogenic variants in this gene present with distinct symptoms however this does not explain isolated phenotype of this patient’s HCM. Despite being found in patients with both sustained monomorphic ventricular tachycardia (SMVT) and DCM, there is currently insufficient evidence to establish a link between the KCNJ2 gene variant and HCM(Kimura et al., 2012; Ortega et al., 2016).

### Dilated cardiomyopathy

In total, patient-18 has variants in the MT-CYB, 16S (MT-RNR2), TTN, DMD, and PLEC genes. Patient-23 has variants in the MT-COI and MT-ND6 genes. Next, patient-24 has two variants in the mitochondrial 16S locus. Additionally, patient-5 has mtDNA variants in the 16S and MT-ND4L genes along with one in BAG3 gene, and patient-15 had only nDNA variants in the JPH2 and PCCA genes latter of which associated with propionicacidemia.

Nonsyndromic DCM is a type of heart condition that is typically inherited in an autosomal dominant way. However, there are some subtypes of this condition, such as JPH2- and TNNI3-related nonsyndromic DCM, that can also be inherited in an autosomal recessive way. When a proband is found to have one of the genetic variants known to cause this condition, it is important to consider the risk of DCM for other family members who may also carry the same genetic variant to explain underlying phenotype (Hershberger & Jordan, 2022). Based on available research, it seems that when there is a loss of function in the JPH2 gene passed down through the autosomal recessive method, it is linked to a severe form of DCM that presents at an early age. On the other hand, when the pathologic variants of JPH2 are inherited in an autosomal dominant manner, it leads to a condition called HCM through a mechanism known as dominant negative effect (Parker et al., 2023). Nevertheless, it may not be sufficient to explain the clinical presentation without conducting a family analysis, but it may also be necessary to evaluate it in co-occurrence JPH2 variant (JPH2(NM_020433.5):c.1879A>G/p.Ile627Val) with a PCCA variant (PCCA(NM_000282.4):c.522G>A/p.Met174Ile) found in in the patient-15. Other studies have shown that certain pathogenic variants can explain the clinical presentation even in combination with other variants in other genes(Riemersma et al., 2017; Sabater-Molina et al., 2016).

For patient-18, it is challenging to determine the role of pathogenic variants in the TTN gene in DCM because a significant number of controls from other studies also have truncating variants in this gene and not all families with DCM have a clear pattern of inheritance of the variants. Additionally, some research have shown that the truncating variants found in individuals with DCM tend to cluster in a specific region of the protein encoded by the TTN gene. So far, missense variants in the TTN gene have not been linked to the development of DCM (Herman et al., 2012; Norton et al., 2013; Roberts et al., 2015). In addition to other alterations in nDNA of this patient, a particular alteration in a specific region of MT-CYB (m.15497G>A), which is highly conserved across different forms and species, has been identified. One study looked at blood and heart tissue samples from people who died suddenly from CM and 151 healthy controls. They found that a specific genetic variation (m.15497G>A) in the MT-CYB gene occurred more frequently in the sudden cardiac death cases. This variation is in a highly conserved part of the gene (89%), and the researchers suggest that it may be associated with reduced energy metabolism (Kobayashi et al., 2011).

Mitochondrial variants at the region whose conservation score bigger than 70% in patient-5 and patient-24 is in MT-RNR2 (m.2294A>G and m.3159AA>T, respectively), in MT-CO1 (m.6261A>G) for patient-23, which are the only changes in these patients. Some studies investigated variants in rRNA and found that some pathogenic variants have been associated with certain diseases in literature(Elson et al., 2015; Gutell et al., 2002; Smith et al., 2013). However, there is currently no tool available to evaluate human-specific MT-RNR1 and MT-RNR2 variants in silico other than methods like those used to study evolutionary ribosomal RNA pathogenicity like HIA (Vila-Sanjurjo et al., 2021). It is suggested that mitochondrial rRNA mutations can affect mitochondrial translation and can cause or modify the phenotype of cardiomyopathy(D. Li et al., 2019; S. Li et al., 2018; Santorelli et al., 1999). On the other hand, it is reported that BAG3 is essential for maintaining mitochondrial homeostasis, and changes in BAG3 expression have an adverse effect on cardiomyocyte function, which can shape the clinic of patient-5 with its mtDNA variants (Tahrir et al., 2017; Wang et al., 2023). A previously reported missense mutation in the MT-CO1 gene has been suggested to have a high likelihood of being associated with myopathy (Zaragoza et al., 2011). It has been noted that MT-CO1 plays a role in Complex IV formation and is important in the context of cardiomyopathies (Huigsloot et al., 2011). Since it is not possible to overlook the cardiac involvement of myopathy, it is not surprising that an effective change in this gene could cause cardiomyopathy. However, ClinVar submissions of these variants are likely benign regarding other mitochondrial diseases, maternal segregation analyses are still needed to clarify the inheritance for DCM.

### Left-ventricular non-compaction

Research has shown that in nearly half of LVNC cases, there are variants found mostly in genes associated with other cardiac-related conditions. However, it is unclear what the definition of the phenotype or the pathogenicity of these variants is (van Waning et al., 2018). Additionally, it has been suggested that isolated LVNC may be a symptom or a phenotype rather than a distinct disease, as few genetic variants was found in cases that also had other cardiomyopathy symptoms (Hershberger et al., 2017; Miller et al., 2017).

For the five cases of LVNC, one pathogenic heterozygous frameshift deletion was found in TRDN gene and other variants were classified as VUS in TSFM, MYH7, TTN, AGL, TRDN, RYR2, RBM20, BAG3, DSP and LARGE2 genes. Patient-21 who has a frameshift deletion in TRDN (TRDN(NM_006073.4):c.423delA/p.Glu142fs) in TRDN gene, classified as pathogenic, but homopolymer nature of the region requires Sanger sequencing to conclude the disease-variant association. Also, this patient has a VUS in RYR2 gene that can contribute to the phenotype because TRDN is a protein that connects the RYR2 gene with some other cardiometabolic proteins and plays a role in regulating the release of calcium in the sarcoplasmic reticulum, which is showed in a functional study (Rebs, 2021).

A frameshift deletion in MYH7 (MYH7(NM_000257.4):c.575delT/p.Ile192fs), a heterozygous stop variant in TSFM (TSFM(NM_001172696.2):c.5C>A/p.Ser2*), and a missense variant in TTN were identified in patient-19. Some biallelic mutations associated with MYH7 have been reported to be associated with LVNC (Kolokotronis et al., 2019). There are also publications linking TSFM gene with mitochondrial cardiomyopathies (Takeda, 2020). Thus, although these mutations are expected to explain the phenotype to some degree, they are classified as VUS. On the other hand, a mutation in TSFM encoding one of the mitochondrial translation prolongation factors has been associated with isolated fatal neonatal HCM in one family (Smeitink et al., 2006). Considering the clinical overlaps of HCM and LVNC (Lin et al., 2021), it is appropriate to re-evaluate these variants after segregation analysis. In addition, two missense variants in TTN and one in RBM20 (RBM20(NM_001134363.3):c.3249G>C/p.Glu1083Asp) were found in patient-26 and classified as VUS. Similarly, many studies have reported that LVNC is caused by mutations in a RBM20. One is the first to report that reduced activity of RBM20 can lead to LVNC due to TTN mis-splicing. Some other studies also revealed that truncating TTN variants are the leading cause for LVNC and introduce variants in RBM20 as a novel etiopathology for LVNC (Anna et al., 2020; Sedaghat-Hamedani, Haas, Zhu, Geier, et al., 2017; Sedaghat-Hamedani, Haas, Zhu, Kayvanpour, et al., 2017). One another study also presented a patient with an early onset DCM carrying a combination of (likely) pathogenic TTN and RBM20 mutations (Gaertner et al., 2021). Additionally, one research found that the splicing of RBM20 target genes is affected in the mutation carrier and revealed RBM20 haploinsufficiency presumably caused by the frameshift mutation in RBM20. The results showed that the same variant in RBM20 can cause DCM, with or without the LVNC phenotype (Sun et al., 2020). Understanding of the variant pathogenicity across the RBM20-coding transcript is needed with segregation analysis for this patient and others who have LVNC, and it’s important for the future management of the disease. For patient-27 who has three heterozygous missense variants each in DSP (DSP(NM_004415.4):c.3226C>T/p.Leu1076Phe), BAG3 (BAG3(NM_004281.4):c.461T>G/p.Val154Gly) and LARGE2 (LARGE2(NM_001300721.2):c.385C>T/p.Leu129Phe), there is no clear explanation for the phenotype without a family study. However, a specific truncating variant was reported in the DSP gene is associated with the development of LVNC and severe early-onset heart failure even though it is typically associated with DCM (Williams et al., 2011). The study findings reveal a range of genetic variations in individuals with LVNC. The various genetic makeup may account for the different clinical presentations observed in patients with LVNC.

In addition to missense TTN variants, three variants that have high conservation scores in MT-CO1 (m.6546C>T), MT-CO3 (m.9948G>A) and MT-RNR1 (100%, 96%, 73%, respectively) were detected in patient-20 whose pedigree is given in **Figure 4C**, and the variant in MT-RNR1 (m.684T>C) could not be found in the databases and seems a novel homoplasmic variant. Patient’s one sibling died of an unknown cause and another sibling also manifests LVNC and some other dysmorphic findings caused by additional *de novo* microdeletion revealed by microarray analysis. In a study that investigates the mitochondrial rRNA encoding the 12S subunit of mitochondrial rRNA, it was stated that LVNC may be associated some variants in this locus. However, they also suggested that the 12S rRNA, MT-RNR1, would have a more smaller effect than other mutations found that changed its conformation (Tang et al., 2010). One study suggests that analyzing mtDNA sequence variants in myocardial tissue samples could provide useful information for understanding the causes of LVNC (Liu et al., 2013). In this study, researchers found that the mtDNA copy number in the myocardium of LVNC patients was lower compared to healthy controls. This lower mtDNA copy number, in combination with morphological abnormalities of mitochondria, suggests that there may be dysfunction of the mitochondria which may play a role in the development of LVNC.

### Restrictive cardiomyopathy

Patients-9 and patient-10, who are cousins on their mother’s side, whose pedigree is shown in **Figure 4C**, were found to have a homozygous pathogenic variant in the TNNC1 gene (TNNC1:p.8A>V). The TNNC1 gene codes for troponin C protein, which is responsible for contractility in the heart muscle and is one of the three members of the complex troponin family. Pathogenic variants of TNNC1 have been associated with DCM and HCM in OMIM and many reported variants in this gene support this association (Parvatiyar et al., 2012; Pinto et al., 2011). Although HCM is typically inherited in an autosomal dominant manner, the presence of homozygous situation in our study, with another mutation in the same gene, causing RCM in a case study presented by Ploski et al., suggests that pathogenic variants in this gene may have an autosomal recessive inheritance pattern for RCM (Ploski et al., 2016). Also, the variability of Ca2+ concentration, which affects myofilament function for each subtype, can create a difference for the expressional variability of cardiomyopathy. For example, high Ca2+ levels during contraction have been associated with hypertrophy in heart muscle, while low levels have been associated with dilation. Hence, the importance of investigating the role of mitochondria, which play a significant role in the storage of cytosolic Ca2+, in terms of the full diagnosis, progression, and treatment of the disease has been approved (Willott et al., 2010).

As stated by the ACMG, interpreting mitochondrial variants is complex and challenging (Richards et al., 2015). Due to the unique characteristics of mitochondrial inheritance, such as heteroplasmy and homoplasmy and specific geographic regions, certain tools should be developed to specifically analyze mtDNA variants. In addition to rare cases, mtDNA, which is transmitted through the maternal line and is different from Mendelian inheritance, plays an important role in the evolutionary process of humans and diseases (Wallace, 2015). Haplogroups, have been the subject of many recent studies, particularly in the context of neurodegenerative diseases and regional/geographic predispositions to certain disorders (Chinnery & Gomez-Duran, 2018; Wallace, 2018). In this study, we investigated possible genetic explanations of pediatric cardiomyopathies by analyzing both nuclear DNA and mitochondrial DNA. Our study is also the first high-throughput sequencing study of mtDNA in Central Region of Anatolia Peninsula (Turkey) that aims to contribute to the country’s mtDNA haplogroup map by interpreting ethnic/regional haplogroup variations. Since it is discussed that genetic predispositions to diseases can be geographically determined(Yusuf et al., 2001), this study conducted in our region presents a promising analysis for promising targeted mitochondrial therapies developed recently(Nissanka & Moraes, 2020). Even though the sample size is not diverse enough to generalize the findings to the larger non-syndromic or isolated pediatric cardiomyopathy population and the study only examined a limited number of genes, this combined analysis could provide insights into the etiology of certain idiopathic cardiovascular diseases.

In addition, considering the large size of TTN and many variants found in this gene, it can be argued that some functional studies should concentrate only on TTN. Furthermore, in this study, we identified 10 novel variants in four genes that could potentially be directly related to cardiomyopathy and two genes that could be indirectly related. Pediatric manifestations with their genetic analyses included in this study will be useful for pediatricians to support the use of genetic testing and for researchers to investigate new genetic factors. In addition, it is essential to evaluate mtDNA variants together with nuclear DNA variants, especially in tissues where bioenergetic pathways are actively involved in the pathology.

## MATERIALS AND METHODS

### Ethical approval

This study was conducted according to the guidelines of the Declaration of Helsinki and approved by the clinical practice ethics committee of a hospital. Informed consent was obtained from the parents or legal guardians of the probands, and each participant provided informed consent. We also followed STROBE guidelines for reporting observational studies(von Elm et al., 2007).

### Diagnosis

All 27 patients were evaluated in pediatric cardiology department between 2018-2020. Transthoracic echocardiogram (TTE) and a 12-lead electrocardiogram (ECG) was used as initial modalities and, 24- to 48-hour ambulatory electrocardiographic monitoring, exercise TTE, intraoperative transesophageal echocardiogram (TEE), cardiac magnetic resonance imaging (MRI) was used when necessary.

Children with positive family history and left ventricular wall thickness more than 2 standard deviations (SD) from the predicted mean and without family history, more than 2.5 SDs were diagnosed with hypertrophic cardiomyopathy (HCM) according to 2020 AHA/ACC guidelines(null et al., 2020). Children with reduced left ventricular ejection fraction (LVEF) (<50%) with enlarged left ventricle volume in the absence of other cardiac findings including coronary artery disease, primary valvular heart disease in addition other systemic and extrinsic reasons including hypothyroidism, toxins and medications were diagnosed with dilated cardiomyopathy (DCM)(Libby et al., 2021). There were only two having restricted cardiomyopathy (RCM) who were children of first-degree cousins. They had reduced ventricular filling with normal/near normal ejection fraction and myocardial thickness and mildly reduced diastolic volumes. These patients were diagnosed with restrictive cardiomyopathy (RCM) in the presence of all these findings according to ESC(Elliott et al., 2007). The left ventricular non-compaction (LVNC) group was diagnosed according to morphological diagnostic criteria stated by Jenni et al; multiple trabeculations in echocardiographic imagining, multiple communicating ventricular cavities and intertrabecular recesses in color Doppler imaging, recesses within the middle and apical side of the ventricle, and a noncompacted to compacted ratio more than 1.4, in addition to the endocardium with two layered constructions(Jenni et al., 2001).

The subjects were then elaboratively evaluated by clinical geneticists to exclude secondary causes of cardiomyopathy, including RASopathies, muscular dystrophies, Barth syndrome and other metabolic disorders. They were evaluated for neurodevelopmental milestones, intellectual capability, and underwent a systemic examination with anthropometric measurements, dysmorphic feature inspection, organomegaly examination, and neurological examination. Perinatal history and pedigree analysis were also assessed, and ophthalmological and auditory examinations were done if needed. Initially, there were 32 participants, but one of them voluntarily withdrew from the study, resulting in a total of 31 participants for the control group. They had no history of heart problems or familial heart and syndromic findings, as confirmed by cardiology and genetics clinic.

### Demographic features

There are a total of 27 patients 12 of whom were female and 15 were male. The mean diagnose age ranged from 0.54 for RCM, 1.41 for DCM to 8.6 for LVNC, with HCM having the second highest mean diagnose age of 8.42 years. The control group included 31 individuals, 21 of whom were female and 10 were male, with a mean age of 10.03 years. The figure (**Figure 2A)** summarizes this feature across phenotypes. Detailed demographic and clinical summary of the study participants can be found in **Supplementary Table S1A.**

### Sample collection and DNA isolation

Approximately 400 µl of peripheral whole-blood samples from 58 individuals were collected into EDTA containing tubes. MagPurix robotic system and its isolation kit were used in accordance with the manufacturer’s instructions. Around 100 µl of total genomic DNA was isolated at a concentration range of 30-100 ng/µl measured by NanoDrop and these samples were stored at −20°C.

### Sequencing and bioinformatic analyses

Library was prepared according to Illumina Trusight Enrichment and clinical exome sequencing was performed in Illumina-NextSeq500 platform. Mitochondrial DNA was subsequently sequenced on IonTorrent-S5 platform. The library preparation was conducted using the Ion AmpliSeq™ Library Kit 2.0 and Precision-ID mtDNA Whole Genome Panel by following the manufacturer’s guide. Sequenced output data (fastq files) were aligned to the related genes of human reference genome (UCSC hg19/GRCh37) by BWA-mem (v.0.7.12) (H. Li, 2013) and quality assessments were performed by using FastQC(S. Andrews, 2010). After quality trimming and filtering the bad reads out, variant calling was performed by GATK (v.4.0)(A. McKenna et al., 2010). For mtDNA variant calling mtDNA_rCRS (NC_012920.1) is used as a reference genome for alignment(R. M. Andrews et al., 1999) and Torrent Variant Caller plugin v.5.2.1.39 was used for variant calling. Confirmation of meaningful variants using Sanger sequencing has not yet been carried out except novel mtDNA variant (m.684T>C).

All variants identified in 137 genes that are associated cardiomyopathy and in mtDNA were obtained in VCF format then filtered according to following criteria: 1) Variants at the exonic regions with AF<0.01 and read depth of at least 20x were included. 2) mtDNA variants were compared to the control group variants, and if they were found in the control group, they were not considered for evaluation.

General data handling was performed with and “R” base functions and (version 4.2.1, [R Core Team 2022]), and R packages; ggplot2 and dplyr for plotting(*Dplyr: A Grammar of Data Manipulation*, 2021; Wickham, 2016). After filters were established for nDNA variants, *in silico* evaluations of the variants were carried out using VEP through different prediction tools and PHACT, which was recently developed(Kuru et al., 2022; McLaren et al., 2016). For mtDNA variants, ‘bam’ files were converted to ‘fasta’ files through MitoSuite, which would give the consensus sequence of the reading. Variant analyses were performed on these ‘fasta’ files through Mitomap-MITOMASTER(Lott et al., 2013). Additionally, these variants identified in the patient group were also investigated *in silico* through PolyPhen2-HumDiv and HumVar, MSeqDR, and HmtVar(Preste et al., 2019; Shen et al., 2018). Population data and conservation scores were obtained through Mitomap. For the mtDNA-phylogenetic tree, variants were evaluated and haplogroups were retrieved from Haplogrep (v.2) using PhyloTree17(Weissensteiner et al., 2016). Chi-squared tests were performed using stats package of R to compare the distribution of distinct haplogroups between patient and control groups and p<0.05 was determined as significant. The mitochondrial solar plot shown in the figure was adapted from Stephen Turner’s github account(*Stephenturner/Solarplot: Mitochondrial Solar Plot*, n.d.).

### Data availability

Sequencing data for cardiomyopathy-associated genes in VCF file except five patients who are not alive will be shared reasonable request to corresponding author. Moreover, aligned mtDNA-seq data for 27 patients and 31 individuals in control group are publicly available upon publication on Sequence Read Archive (SRA) under BioProject Number PRJNA1028644 on NCBI. Notably, unfiltered variants for both mtDNA and nDNA are provided in **Supplementary Table S1B, Table S1C, and Table S1D** along with the information of 137 genes.

## Data Availability

All data produced will be available online upon publication under BioProject Number PRJNA102864 on NCBI.

## ACKNOWLEDGEMENTS & FUNDING

The designed study has been supported by scientific grant programs within the university under the supervision of Dr. Ebru Erzurumluoglu Gökalp to support Mehmet Arda Temena’s master thesis.

## SUPPLEMENTARY FILES

### Supplementary Figures

**Supplementary Figure S1.** Summary of nDNA Variants Grouped by Genes and Genotypes.

**Supplementary Figure S2.** Summary of VUS and Pathogenic nDNA Variants Grouped by Genes, Genotypes, and Cardiomyopathy Subtypes

**Supplementary Figure S3.** All mtDNA Variants Identified in Patients.

**Supplementary Figure S4.** All mtDNA Variants Identified in Control Group.

**Supplementary Figure S5.** Filtered mtDNA Variants Grouped by Genes, Genotypes, and Cardiomyopathy Subtypes

**Supplementary Table S1.** Detailed information of patients and all 137 nDNA genes and variants

**Supplementary Table S2.** Detailed haplogroup information and mtDNA variants.

## AUTHOR CONTRIBUTIONS

MAT and EEG conceptualized and designed the project. Funding acquisition was done by EEG and OC. MAT and DC performed all wet-lab procedures. MAT performed bioinformatics analyses and visualized the data. MAT, EEG and ES interpreted the results. PG and HK were involved in diagnosing the patients. MAT, EEG, and ES provided discussion. BDA, SA and OC supervised the project. MAT wrote the manuscript with final approval of all authors.

