## Supplementary Figures for "Uncovering the Dual Role of Mitochondrial and Nuclear DNA Variants in Pediatric Cardiomyopathies"

**Supplementary Figure S1.** Summary of nDNA Variants Grouped by Genes and Genotypes

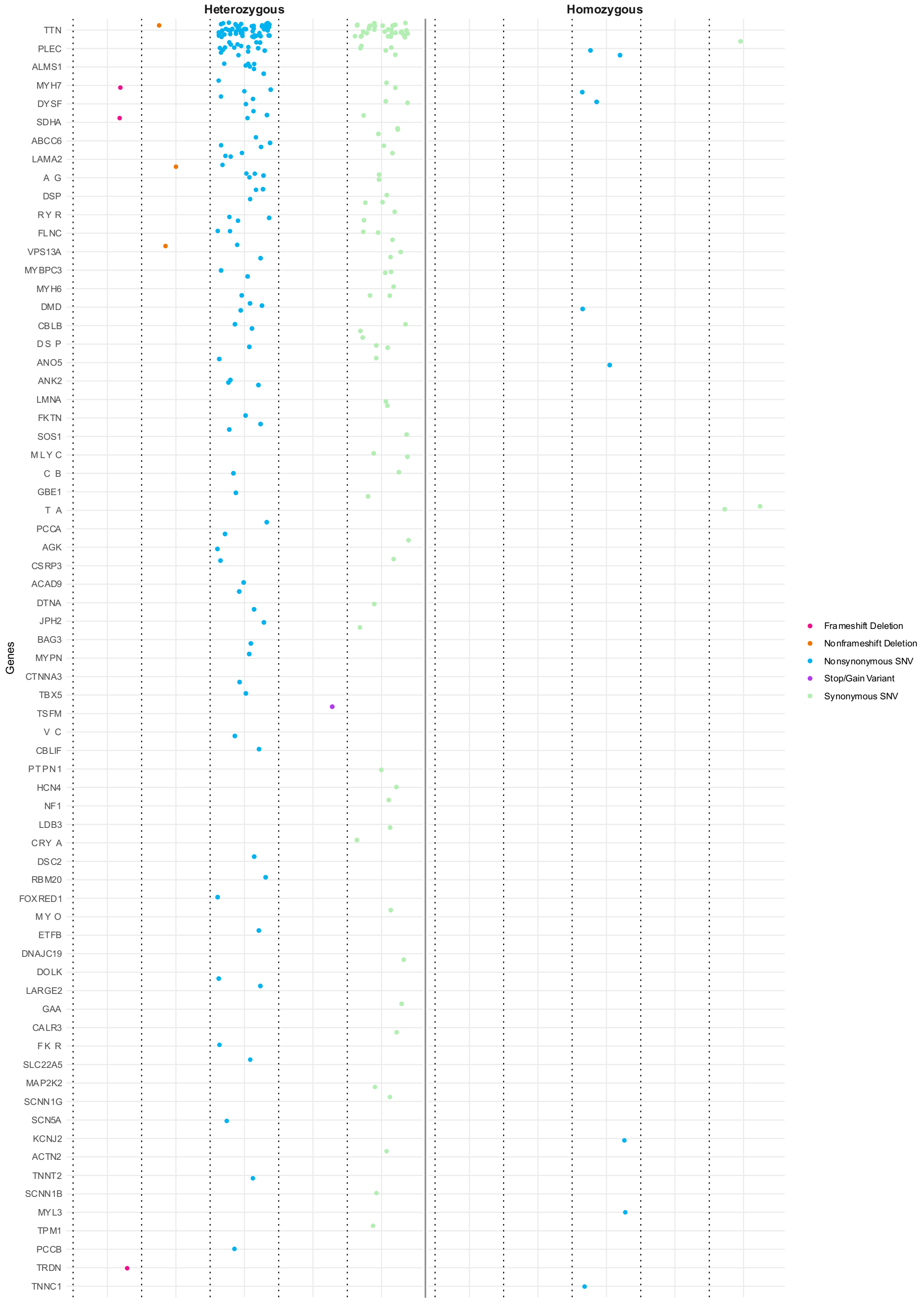

**Supplementary Figure S2.** Summary of VUS and Pathogenic nDNA Variants Grouped by Genes, Genotypes, and Cardiomyopathy Subtypes

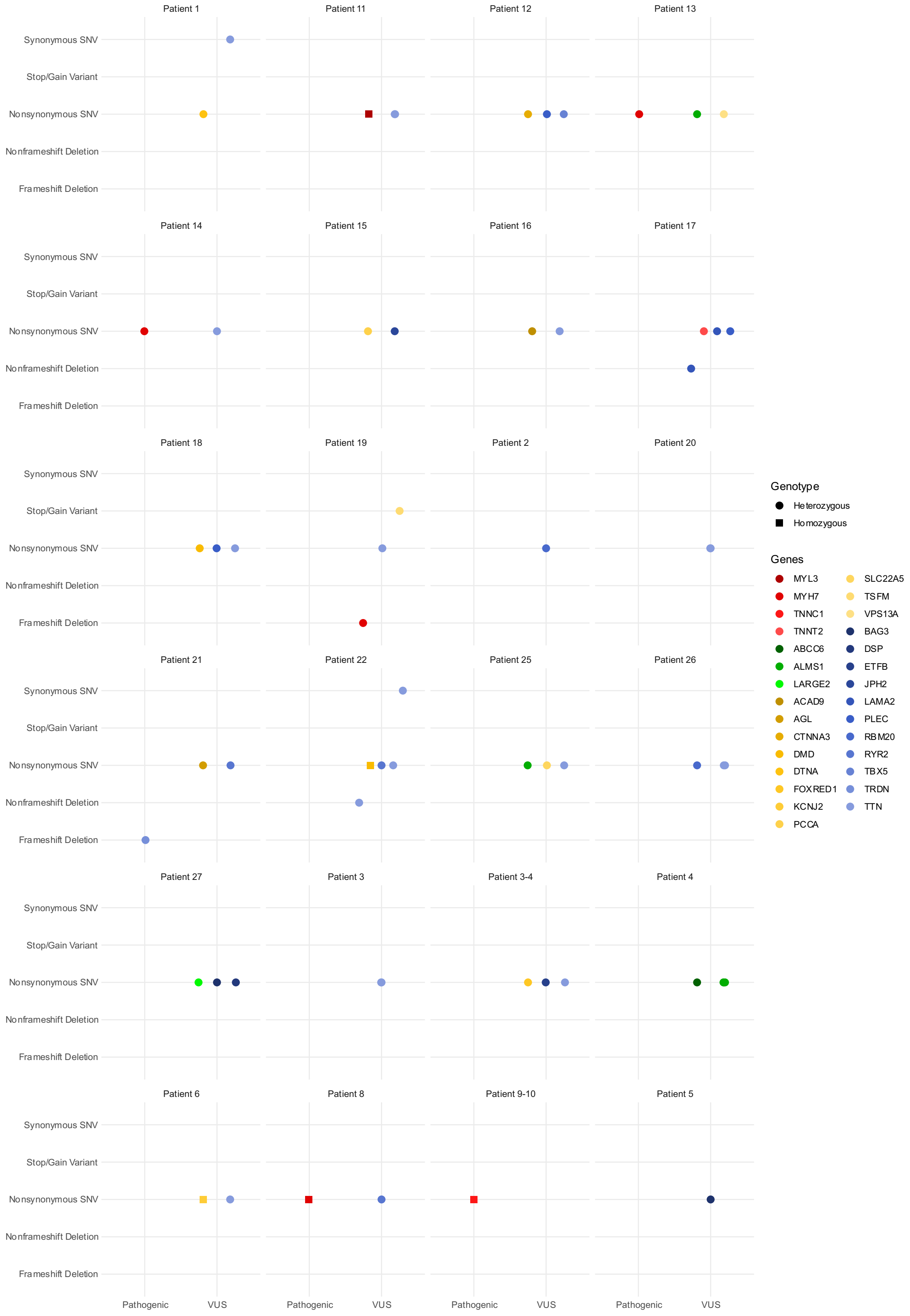

**Supplementary Figure S3.** All mtDNA Variants Identified in Patients.

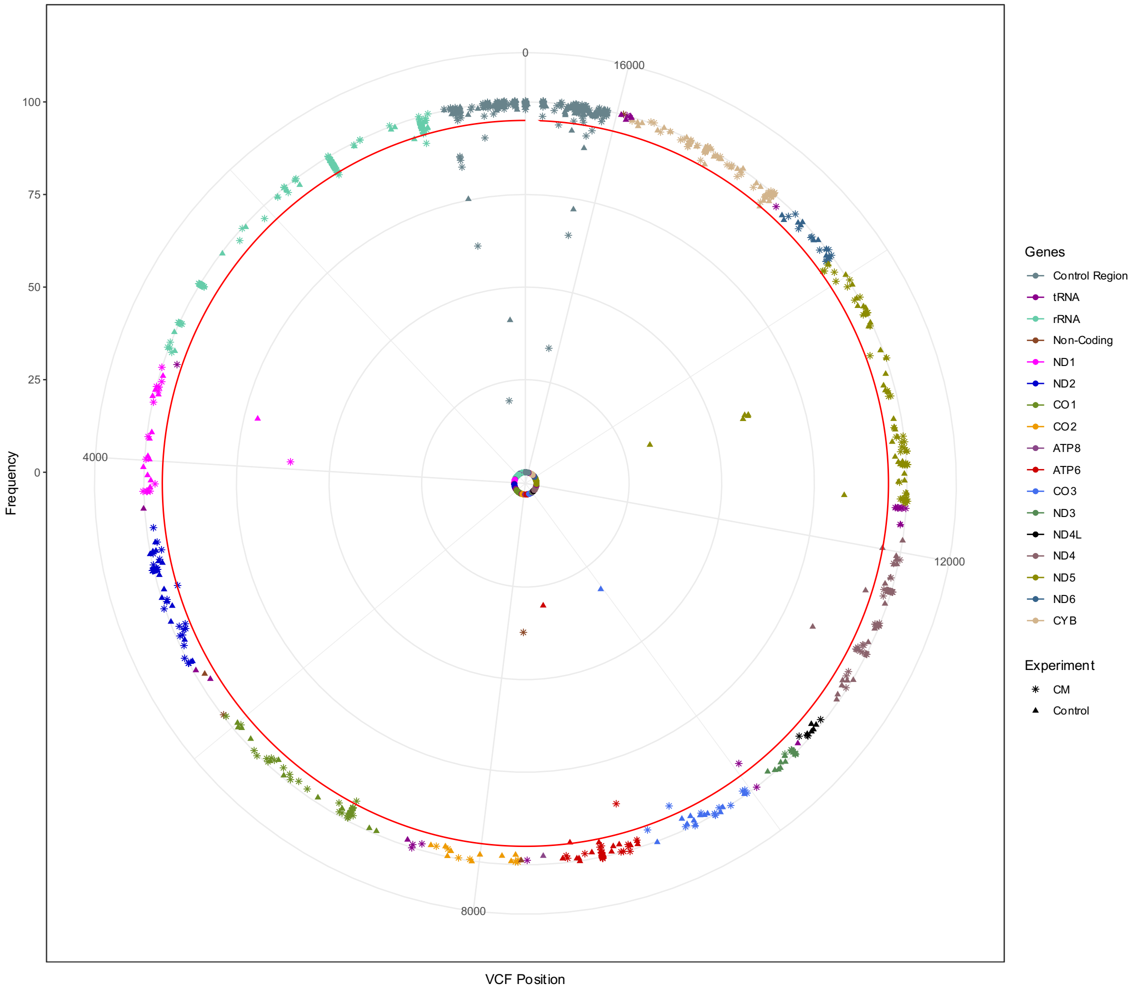

**Supplementary Figure S4.** All mtDNA Variants Identified in Control Group

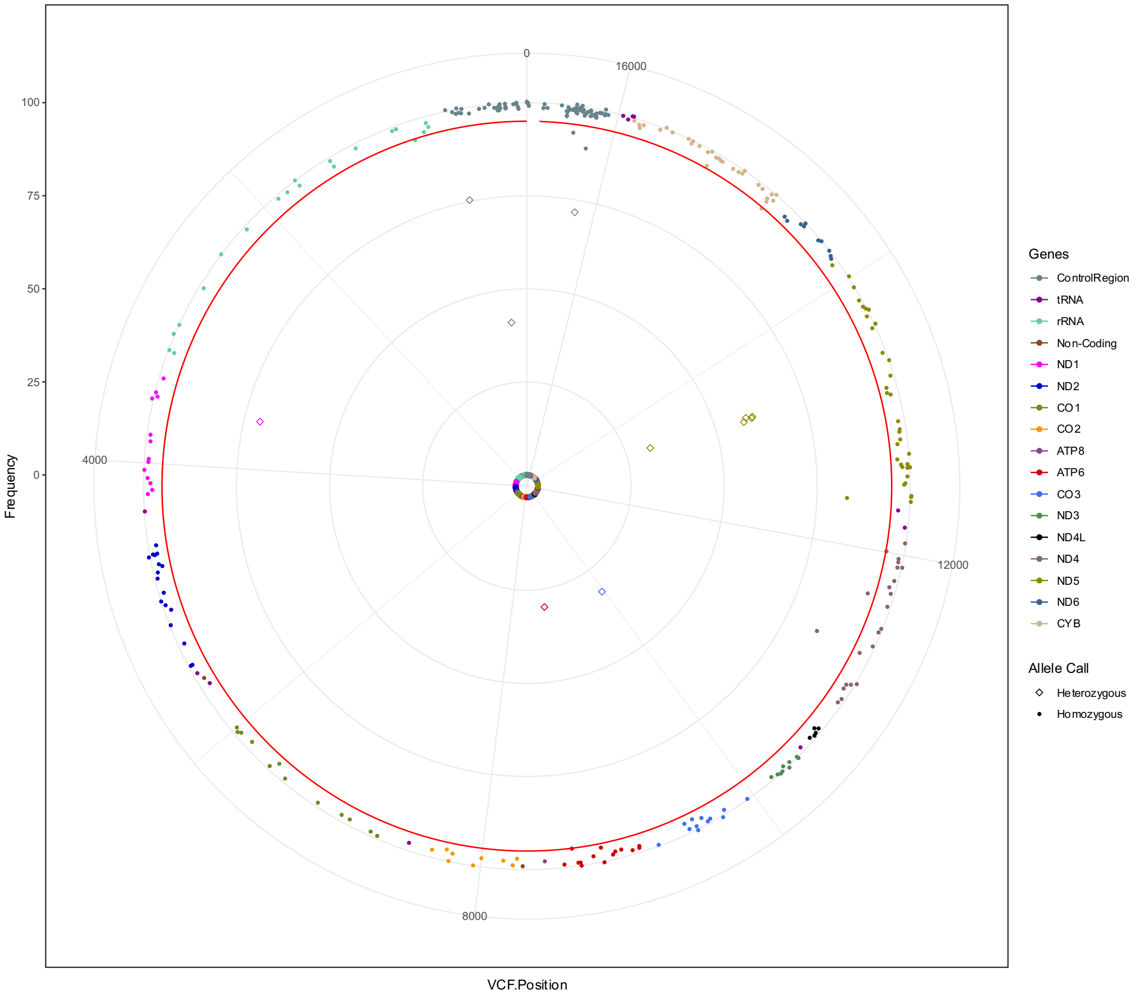

**Supplementary Figure S5.** Filtered mtDNA Variants Grouped by Genes, Genotypes, and Cardiomyopathy Subtypes

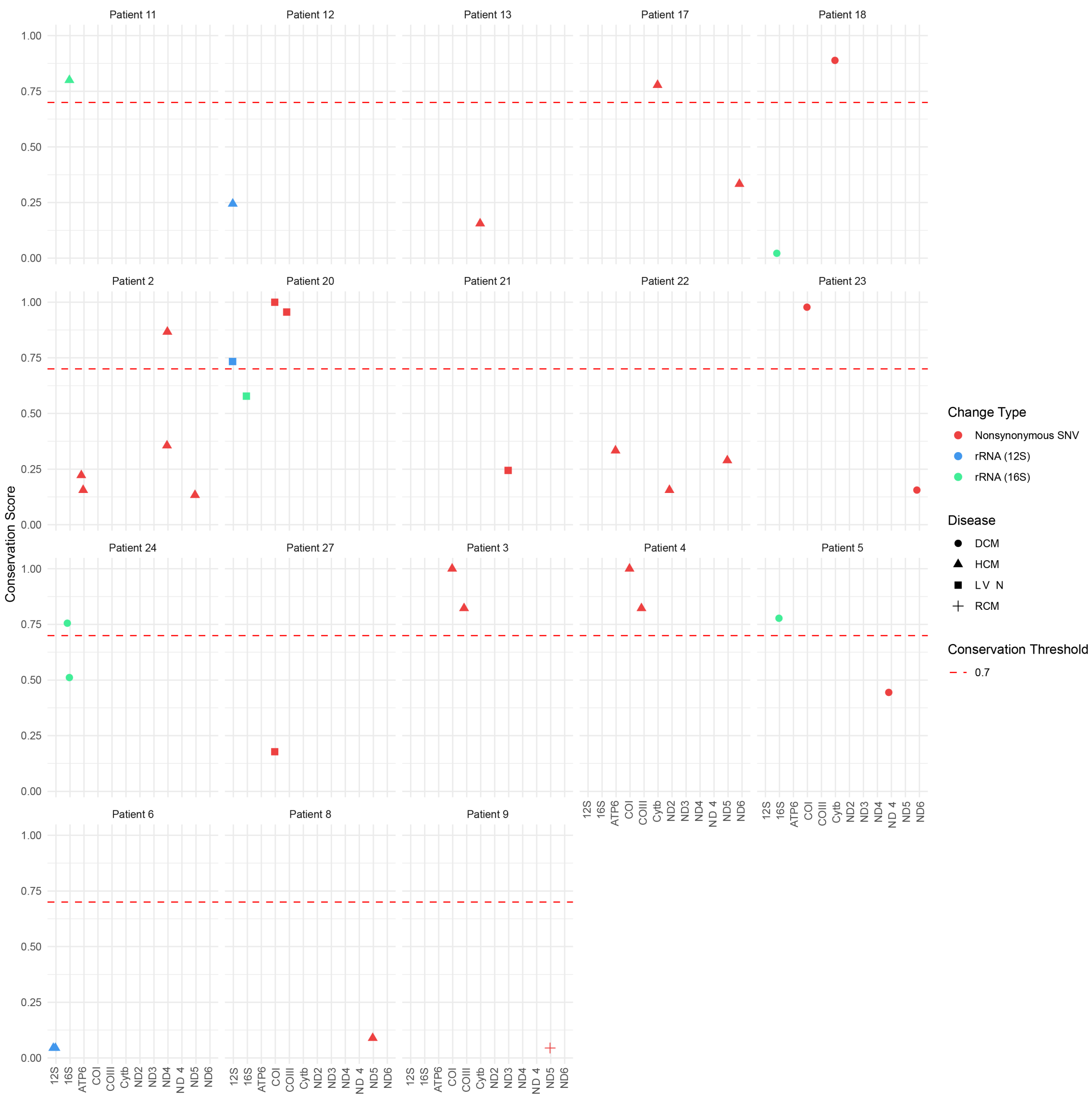
